# Examining inequality in healthcare utilization during pandemic disruptions

**DOI:** 10.1101/2025.04.24.25326361

**Authors:** Jason Lian, Sen Pei, Jianxi Gao, Lu Zhong

## Abstract

**Importance:** Health crisis like recent COVID-19 pandemic has continuously disrupted healthcare utilization, particularly among non-COVID-19 patients. It remains unclear whether these disruptions were experienced equally across populations or if they disproportionately impacted specific demographic groups.

**Objective:** To assess inequality in healthcare utilization during the COVID-19 pandemic, stratified by demographic characteristics (age, race and ethnicity, income, and education) and medical specialties (emergency medicine, anesthesiology, and cardiology) across U.S. states.

**Design, Setting, and Participants:** This retrospective observational study analyzed electronic health records (EHRs) from millions of anonymized patients across U.S. states between 2019 and 2022. The Gini coefficient was used to quantify disparities in healthcare utilization. A regression discontinuity design was employed to assess the effect of declining telehealth usage on inequality.

**Main Outcomes and Measures:** Temporal inequality in healthcare utilization by demographic group and specialty, measured monthly using the Gini coefficient.

**Results:** Compared with the pre-pandemic period (2019), there was an approximate 45% decline in patient visits during the pandemic (2020–2022). Inequality in healthcare utilization increased consistently across demographic groups, with Gini coefficients rising by 14.5% (age), 2.3% (race and ethnicity), 7.0% (income), and 4.8% (education). States that maintained high levels of telehealth service usage experienced smaller increases in inequality.

**Conclusions and Relevance:** The pandemics significantly exacerbated disparities in healthcare utilization among non-COVID-19 patients. These findings underscore the importance of sustaining adaptive healthcare delivery strategies, such as telehealth, to mitigate systemic inequities during long-term public health emergencies.

## Introduction

Natural disasters, including climate change-induced events, environmental pollution, and global pandemics, are recurrent and prolonged crises that starkly exacerbate existing health inequalities within communities^1–6^. The COVID-19 pandemic, for example, has led to approximately 1.2 million deaths in the US, with Black, Hispanic, and Asian populations experiencing significantly higher rates of infection and death compared to White populations^7–11^. Simultaneously, the pandemic has disrupted non-COVID-19 healthcare services^12–15^. Evidence has shown that about 9.4 million cancer screenings and treatments were either delayed or canceled^16^. These declines, spurred by public fears of infection during healthcare visits, lockdown measures, and the social demographic characteristics of patients, likely worsened pre-existing health disparities^15,17,18^. For instance, individuals from vulnerable socioeconomic backgrounds or those residing in areas with limited healthcare resources may have faced more significant disruptions^7^. Yet, it is unclear whether the declines were equally experienced across all patients or if they disproportionately impacted certain demographics.

Numerous studies have focused on health inequality during the pandemic, primarily examining aspects such as exposure risk, disease incidence, hospitalizations, and mortality rates across various categories like age, disability, gender, race and ethnicity, sexuality, occupation, and socioeconomic status^7–10,19–21^. However, there has been less investigation into inequalities in non-COVID-19 healthcare utilization^17,22^ Furthermore, these studies often rely on static measures, failing to consider the temporal dynamics of inequality that may have worsened—or been alleviated—by adaptive responses (i.e., telehealth usage) within healthcare systems during the pandemic^23–26^. Neglecting non-COVID-19 healthcare and failing to track the changes in equality could impede the development of effective, sustainable strategies to mitigate health disparities during prolonged crises^27–29^.

This study aims to address the gaps by assessing the decline in non-COVID-19 healthcare utilization in the United States and tracking the change of inequalities during the pandemic. By analyzing millions of electronic medical records (EMRs) from 2019 to 2022 across various US states, we apply the Gini coefficient^30,31^ to measure monthly inequality among patient demographics, such as age, race and ethnicity, income, and education level. We stratify this inequality by different states and medical specialties to pinpoint vulnerable geographical locations and specialties. Additionally, we conduct the analysis to assess the impact of telehealth usage, i.e., an alternative to in-person healthcare access, on reducing inequality. This research, grounded in real-world data, provides insights into how inequalities in healthcare evolved during the pandemic and offers valuable information to healthcare providers on how to better prepare for and respond to future health crises.

## Methods

### Dataset

The EMR dataset we employ, known as the Healthjump dataset, is sourced from the COVID-19 Research Database^32^. Healthjump operates as a data integration platform that compiles electronic medical records and practice management systems. It contains information on over 33 million patients and 1 million healthcare providers. It features a comprehensive range of patient data, such as diagnoses, procedures, encounters, and medical histories, all sourced from the participating entities within the Healthjump network. It encompasses socio-demographic attributes of patients, including age, race, gender, and more.

Additional demographic information, such as education and income levels, is incorporated by matching patient records based on their home locations with data from the American Community Survey (ACS) provided by the U.S. Census Bureau^33^. Unlike age, race, and ethnicity—which are directly recorded in the EMR dataset—education and income data are only available at the ZIP code level. To estimate these demographics for patients, the proportion of the ZIP code population belonging to specific sociodemographic groups is used as a proxy for the likelihood that patients belong to those groups. These probabilities are multiplied by the total number of patients in each ZIP code to estimate the number of patients with various education and income levels. Summing these counts across all ZIP codes within a state yields the total number of patients in each sociodemographic group.

The telehealth usage dataset is obtained from Data.Medicaid.Gov^34^ and includes monthly counts of services delivered via telehealth, such as live audio-video, remote patient monitoring, store-and-forward, and other telehealth modalities, to Medicaid and CHIP beneficiaries, categorized by state.

### Medical specialties

The medical specialties that patients seek are identified through ICD-10 codes within their diagnoses ^35^. Based on this classification (see Table S1) we analyzed the top three medical specialties in our dataset with the most records. They are emergency medicine, anesthesiology, and cardiology.

### Patient Demographic groups

We categorize patients based on specific demographic characteristics including age, race and ethnicity, gender, income level, and education level. Age groups are defined as Children (≥18 years), Young Adults (18-44 years), Middle-aged Adults (45-65 years), and Seniors (>65 years). Racial categories include Hispanic, White, Black, and Asian. Income levels, measured in annual income in dollars, are segmented into ≤10k, 10k-50k, 50k-100k, 00k-200k, and ≥200k. Education levels are classified as Less than High School Graduate, High School Graduate, Bachelor’s Degree, and Higher Degree.

### Data Preprocessing

Data Preprocessing Steps:

a. Population Selection: Unique patient IDs recorded between 2017 and 2019 are extracted to establish a constant population base, ensuring that changes in population size do not affect the results.
b. State Filtering: States with patient data representing less than 0.1% of the state population are excluded to eliminate states with insufficient data. 44 states are selected.
c. Specialty Selection: The top three medical specialties with the highest number of patients are identified for focused analysis.

### Measure of inequality

Assessing health inequality is crucial for guiding targeted initiatives to reduce them^31^. Tracking changes in inequality over time can reveal progress or setbacks in efforts to mitigate it. We use the Gini coefficient^30^ to measure inequality within each demographic attribute, as follows,

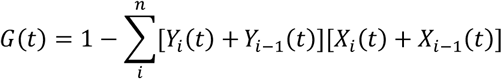

where *X*_*i*_(*t*) is the cumulative percentage of population after including group *i* at time *t*. *Y*_*i*_(*t*) is the percentage of patient visits within the EMR dataset after including group *i* at time *t*. *N*is the total number of groups. We use Gini coefficients to measure monthly inequality within each demographic factor.

### Test impact of declining telehealth usage

As the COVID-19 pandemic progressed, telehealth usage fluctuated, and disparities in access appeared to increase. To assess the impact of the decline in telehealth usage after April 2021, we first need excludes the confounding effects of the pandemic itself. To do this, we use a K-means clustering algorithm^36^ to group US states that had similar levels of pre-pandemic inequality (measured across demographical categories such as age, race, ethnicity, education, and incomes) well as similar baseline levels of telehealth usage. This clustering allows us to compare states with comparable structural characteristics. Within each group, we then apply the linear regression to estimate the impact of COVID-19 infection rates on inequality in the first three month of pandemic, that 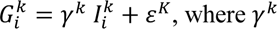 is the group-specific effect represented by for group *k*.

By assuming that the impact of COVID-19 infections (*γ*^k^) keep constant, we utilize the regression discontinuity design^37^ to test the impact of declining telehealth usages, the inequality at state *i* within group *k* at time *t* is modeled as follows,

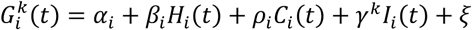

where 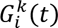 is the Gini coefficient of state *i* within group *k* at time *t*. *C*_i_(*t*) represents the telehealth usage at state *i* at time *t*, and *X*_i_(*t*) is binary indicator equal to 1 if *C*_i_(*t*)exceeds a predetermined threshold based on data, and 0 otherwise. The threshold is defined as the average telehealth usage of three months prior to April 2021 at each state. The variable *I*_i_(*t*) denotes COVID-19 infection rate, and *γ*^k^ the effect of infections on inequality within group *k* and *ξ* is the error term. If *ρ*_i_<0, if means that decline the telehealth usage, lead to increase of inequality in healthcare utilization.

## Results

### Temporal inequality during pandemic disruptions

We calculate the monthly visits of patients who seek healthcare services within the Healthjump EMR datasets as the healthcare utilization. By establishing the average visits over months in 2019 as the baseline, we examine changes in healthcare utilization across US states from 2020 to 2022. As depicted in Fig. 1 and Fig. S1, healthcare utilization sharply declined beginning in March 2020, coinciding with the start of lockdown measures, and reached its low point around August 2020. Subsequently, patient visits showed fluctuating reductions through the end of 2022, with around 45% losses in total in Fig. S1. The drop in healthcare utilization was broadly experienced across specialties. In Fig. S1. the service decline was most pronounced in emergency medicine, with a decrease of 59%, compared to anesthesiology, which experienced a 41% reduction, and cardiology, which saw a 55.4% decrease.

**Figure 1.**
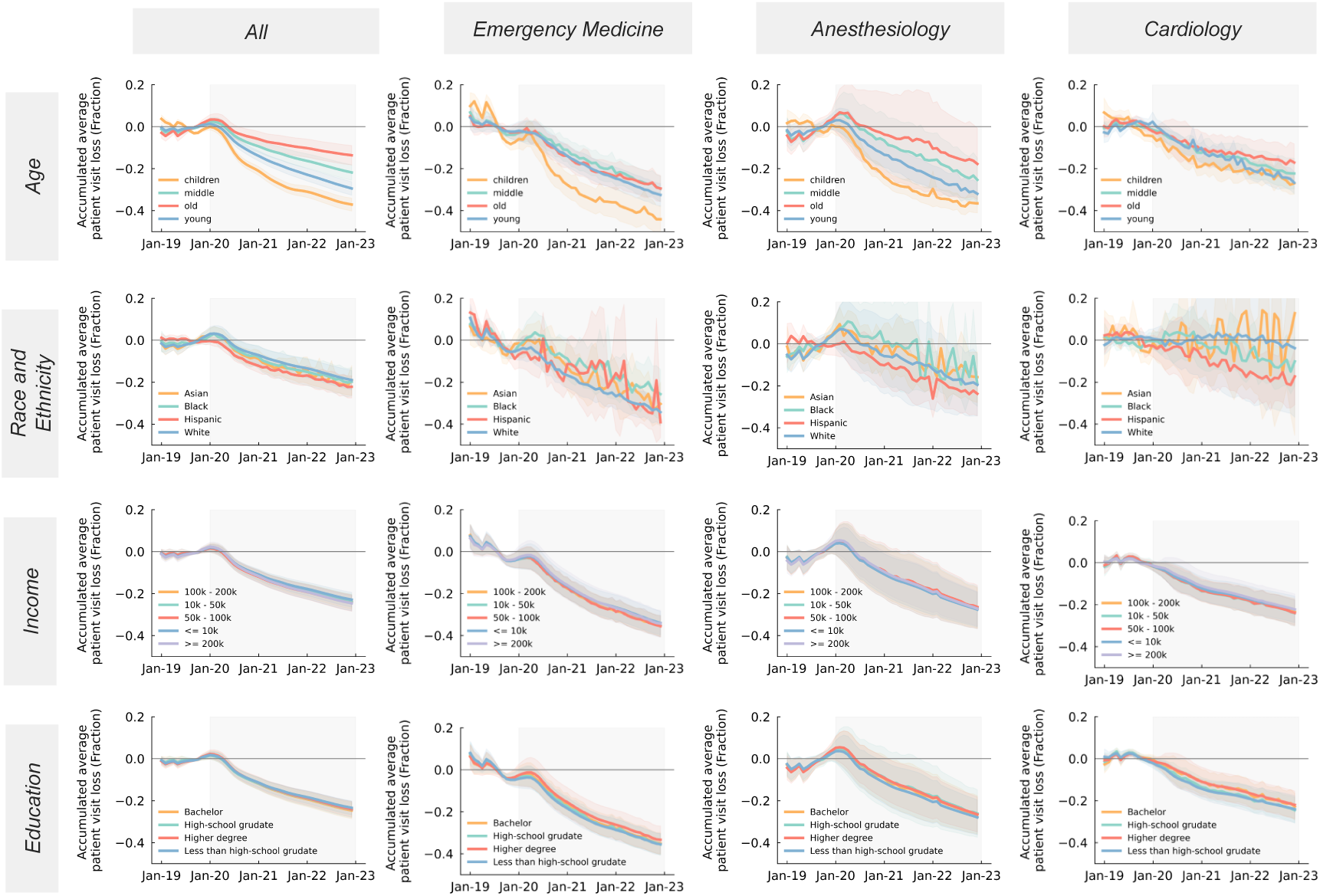
Temporal patient visit loss from 2019 to 2022. Compared to the pre-pandemic period (2019), the cumulative average of patient visits during the pandemic period (2020-2023) have continued to decline across various medical specialties.

Figure 1 illustrates the cumulative average loss in patient visits, segmented by demographic categories including age, race and ethnicity, income, and education level. The results show that the elderly demographic (aged 65 and older) exhibited the smallest decline in patient visits. This was followed, in order of decreasing magnitude, by middle-aged adults (45-65 years), younger adults (18-44 years), and children (18 years and younger). Racial disparities in the reduction of patient visits were relatively modest; however, the Hispanic population experienced a notably higher reduction compared to other racial groups. Individuals with high incomes (exceeding 200,000 annually) experienced fewer reductions in visit frequency compared to those with lower incomes (less than 10,000 a year). With respect to educational attainment, individuals with degrees higher than a bachelor’s experienced fewer reductions in visits, particularly in specialized fields such as emergency medicine, anesthesiology, and cardiology.

We utilize the Gini coefficient^31,38^ to analyze the distribution of healthcare utilization among patient visits in our dataset, relative to the population of each state. Higher Gini coefficient (ranging from 0 to 1) indicates greater inequality. Table 1 displays the Gini coefficients across different demographics during pre-pandemic (2019) and during the pandemic period (2020-2022). Using the average Gini coefficient from 2019 as a baseline, we also present the changes illustrated in Fig. 2. Age demographics exhibit the largest inequality, with a Gini coefficient of 0.357 before the pandemic, which increased to 0.409 during the pandemic—a 14.5% rise. Racial and ethnic inequalities were initially lower, around 0.150 pre-pandemic, with a slight increase to 0.153, indicating a minor change. Income and education disparities were modest, but saw significant increases during the pandemic, rising by 7% and 4.8%, respectively. Among medical specialties, emergency medicine maintained the lowest inequality compared to anesthesiology and cardiology (Table 1). Notably, there were significant increases in inequality within emergency medicine and anesthesiology across age groups, and an increase in anesthesiology and cardiology related to income. Unlike other demographic categories, race and ethnicity did not show increases in 2020 and 2021 but did start to show an increase in 2022, which may be attributed to the long-term impacts of COVID-19.

**Figure 2.**
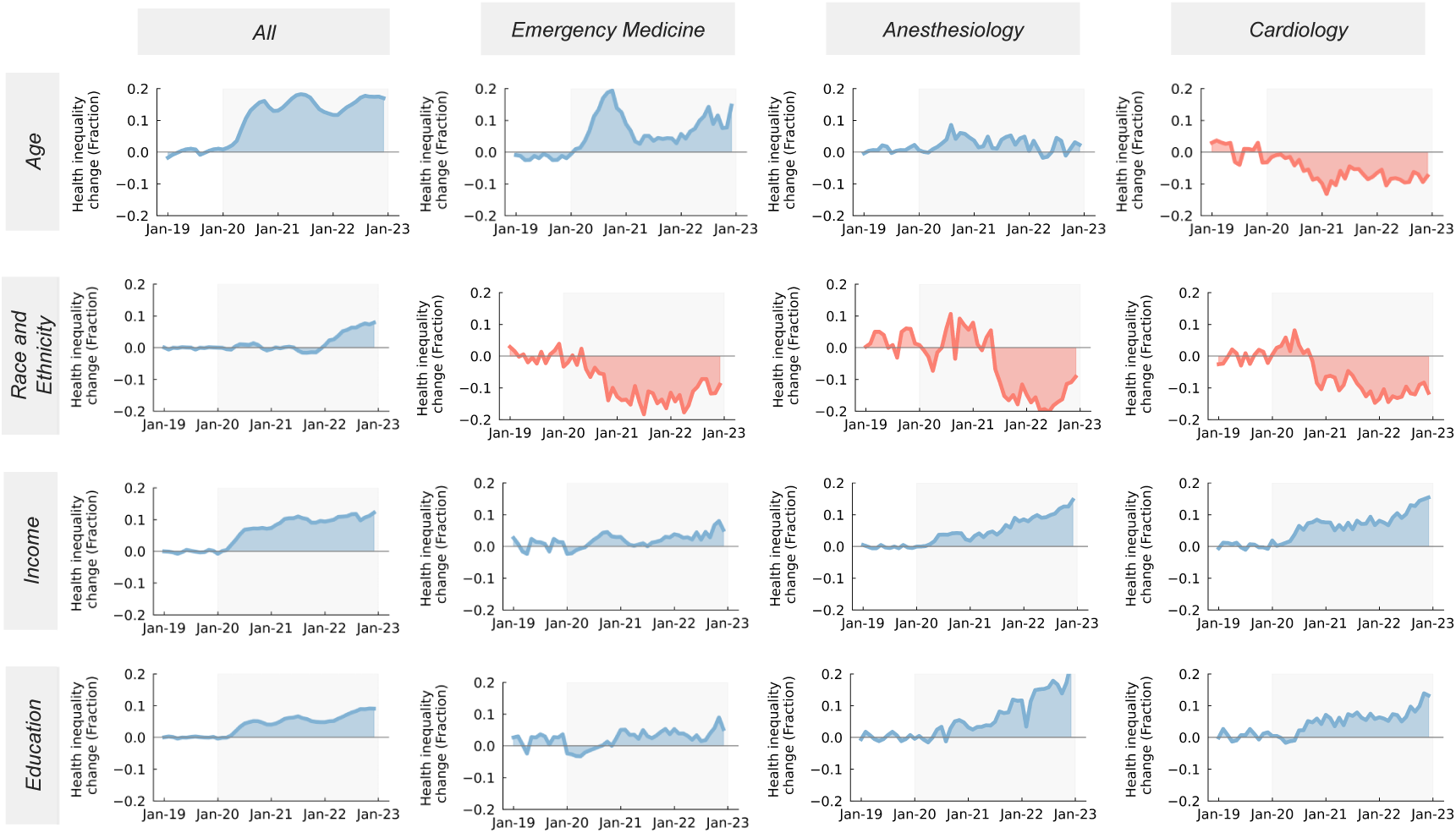
Temporal inequality changes in healthcare utilization from 2019 to 2022. Compared to the pre-pandemic period (2019), the inequality, measured by the Gini coefficient, fluctuated during the pandemic. The inequality for “All” specialties continues to increase for patient visits across all socio-demographic groups. The increase in inequality from 2020 to 2022 is represented in blue, while the decrease is shown in red.

**Table 1.** Inequality in healthcare utilization. The differences between the pre-pandemic and during-pandemic period were tested using one-sided t-tests and were considered significant when the P-value was less than 0.05.

| Characteristics | Pre-pandemic | During-pandemic | Difference |  |
| --- | --- | --- | --- | --- |
|  | 2019<br>Mean [Range] | 2020-2022<br>Mean [Range] | Mean Difference<br>[P-value] | Mean Difference<br>Percent |
| **Age** |  |  |  |  |
| All services | 0.357 [0.349-0.366] | 0.409 [0.404-0.414] | **0.052 [P=0.001]** | 14.5% |
| Emergency Medicine | 0.262 [0.253-0.271] | 0.285 [0.278-0.292] | **0.023[P=0.001]** | 8.9% |
| Anesthesiology care | 0.431 [0.421-0.44] | 0.447 [0.44-0.454] | **0.016 [P=0.009]** | 3.8% |
| Cardiology | 0.494 [0.48- 0.508] | 0.481 [0.471-0.49] | -0.013 [P=0.928] | -2.7% |
| **Race and Ethnicity** |  |  |  |  |
| All services | 0.150 [0.141-0.158] | 0.153 [0.148-0.158] | 0.003 [P=0.25] | 2.3% |
| Emergency Medicine | 0.182 [0.169-0.196] | 0.176 [0.167-0.184] | -0.007 [P=0.785] | -3.6% |
| Anesthesiology care | 0.19 [0.177-0.204] | 0.182 [0.174- 0.19] | -0.008 [P=0.852] | -4.4% |
| Cardiology | 0.182 [0.168-0.196] | 0.167 [0.16-0.175] | -0.015 [P=0.974] | -8.2% |
| **Income** |  |  |  |  |
| All services | 0.059 [0.056-0.063] | 0.064 [0.062-0.066] | **0.004[P=0.026]** | 7.0% |
| Emergency Medicine | 0.078 [0.074-0.082] | 00.08 [0.077-0.083] | 00.002 [P=0.236] | 2.5% |
| Anesthesiology care | 0.079 [0.074-0.083] | 0.083 [0.081-0.086] | **0.005 [P=0.046]** | 5.8% |
| Cardiology | 0.082 [0.077-0.086] | 0.086 [0.084-0.089] | **0.005 [P=0.042]** | 5.5% |
| **Education** |  |  |  |  |
| All services | 0.073 [0.069-0.076] | 0.076 [0.074-0.078] | **0.004 [P=0.049]** | 4.8% |
| Emergency Medicine | 0.09 [0.085-0.095] | 0.091 [0.089-0.094] | 0.001 [P= 0.299] | 1.6% |
| Anesthesiology care | 0.085 [0.081-0.090] | 0.088 [0.086-0.091] | 0.003 [P= 0.139] | 3.5% |
| Cardiology | 0.1 [0.095-0.105] | 0.102 [0.1-0.105] | 0.003 [P=0.173] | 2.7% |

### Health Inequality across states

We also examined the variation in healthcare utilization inequality across different states. Figure 3a presents four dimensions of inequality, each corresponding to different demographic categories. Within our dataset, most states exhibit higher inequality among age groups, which is understandable given that different age groups have been impacted by the COVID-19 pandemic to varying degrees and require different levels of medical care. For race and ethnicity, states such as New York, Massachusetts, Pennsylvania, and Connecticut show higher levels of inequality in healthcare utilization. These states also exhibit greater income-related inequalities. Overall, most states display greater inequality during the pandemic compared to pre-pandemic levels. Figure 2b provides an average of the inequalities across all states for all services.

**Figure 3.**
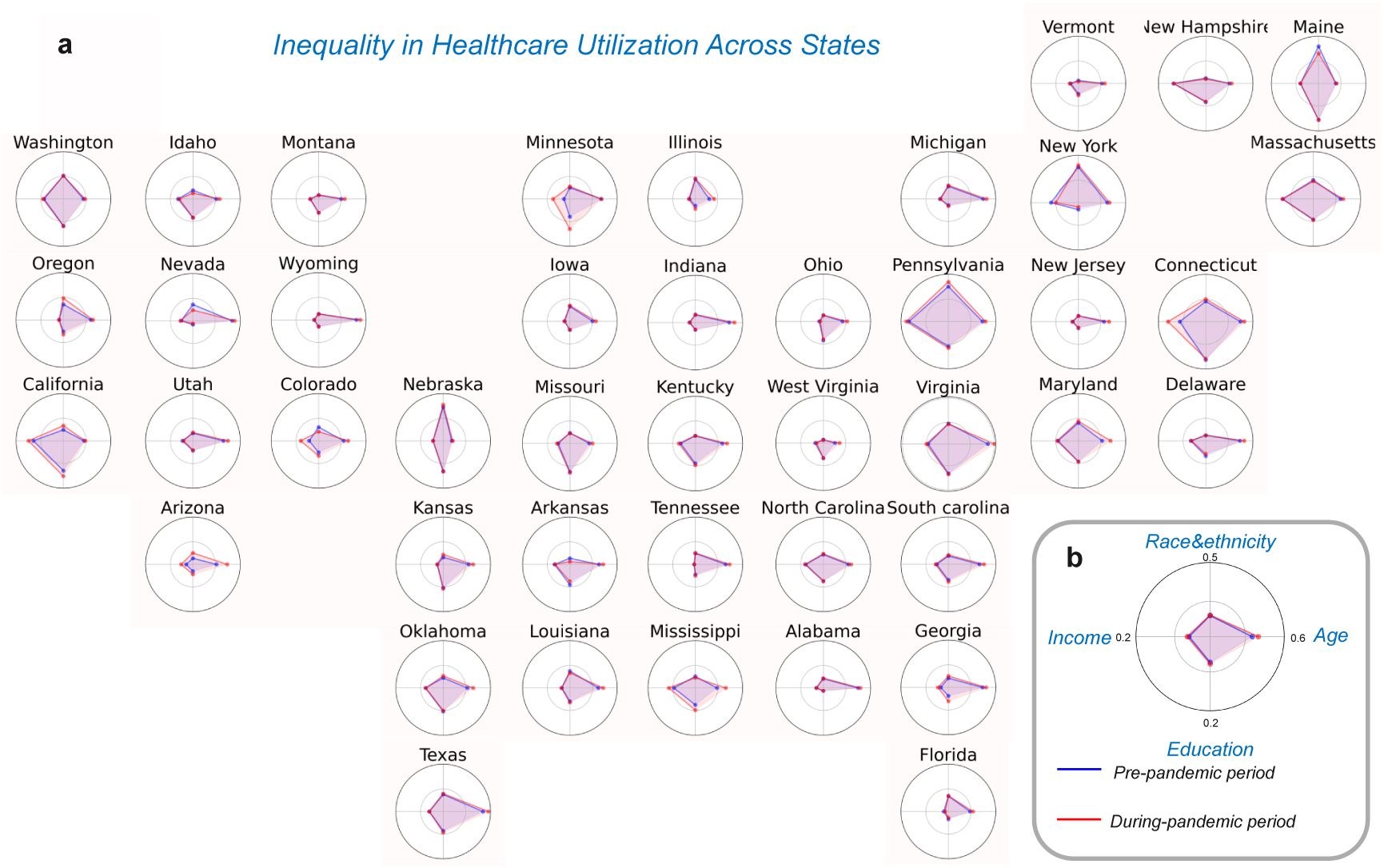
Inequality in healthcare utilization across states. We represent the Gini coefficient among groups of different ages, races and ethnicities, income levels, and education levels as four dimensions in a radar plot and the scale at each dimension is normalized. (a) Inequality in each state. (b) The average inequality across states.

### Association with adaptive actions: Telehealth use

Telehealth, which leverages technology such as phone calls to deliver healthcare services remotely, offers an alternative to traditional in-person healthcare, as recorded in the EMR dataset^25,26^. As the COVID-19 pandemic progressed, congress significantly relaxed Medicare restrictions and expanded reimbursements, leading to a surge in telehealth usage^25^. As illustrated in Fig. 4 a, telemedicine encounters rose sharply in the first three months of the pandemic, increasing from 0.03% to approximately 4% of all healthcare interactions. Subsequently, telehealth utilization began to decrease from April 2021, exhibiting a gradual decline to approximately 1.5% by the end of 2022. Concurrently, the inequality measure (Gini coefficient) demonstrated fluctuations and a subsequent increase. These notable changes prompt an investigation into whether the reduction in telehealth usage contributed to the increase in health inequality from April 2021 to the end of 2022.

**Figure 4.**
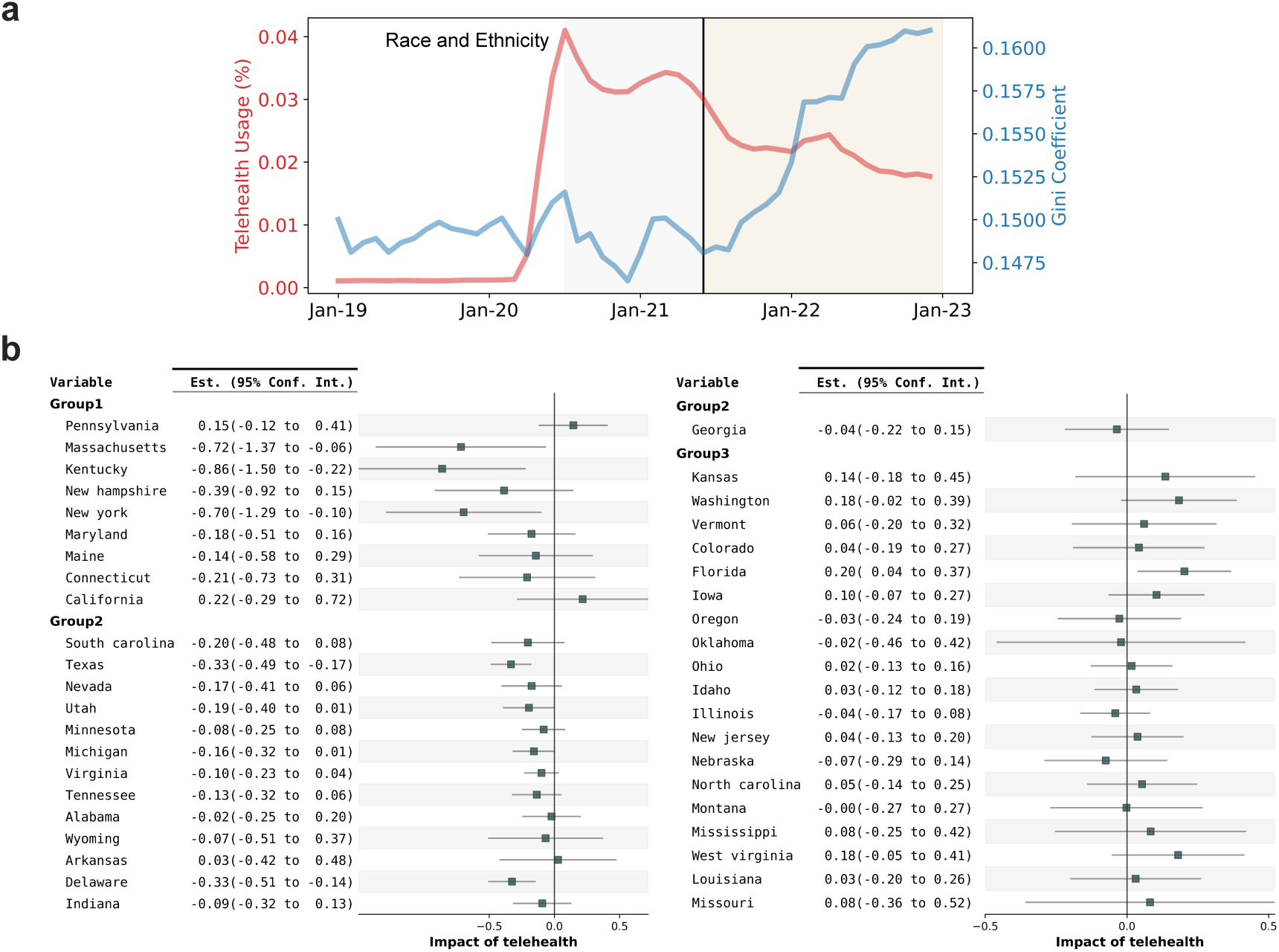
Comparison between inequality in healthcare utilization and telehealth usage. With race and ethnicity group as the example, (a) temporal telehealth usage, in comparison with the inequality across states. We employ regression discontinuity to assess the impact of telehealth usage decline since April 2021 (indicated by the vertical line in (a)) by grouping states based on pre-pandemic inequality levels. (b) shows the effects of telehealth decline across various states. The negative outcomes indicate an increase in inequality correlating with the reduction in telehealth usage.

To exclude the effects of the pandemic, we employed the K-means algorithm to cluster states into groups based on similar levels of pre-pandemic inequality and telehealth usage. This clustering allows us to examine the impact of COVID-19 prevalence on increasing inequality for similar states. Assuming the impact of COVID-19 prevalence on inequality remained steady from 2020 to 2022, we applied the regression discontinuity analysis to evaluate the impact of declining telehealth usage across states, using three-month average telehealth engagement levels before April 2021 as the threshold. Our results, with inequality among race and ethnicity category as the example in Figure 4b, show a negative impact in 25 of the 44 analyzed states (72.1%), meaning the impact of declining telehealth usage in increasing inequality. More pronounced negative effects are observed in State Group 1, marked by high pre-existing inequality and extensive telehealth usage, with average effects of −0.31±0.3. State Group 2, characterized by high inequality but moderate telehealth usage, showed average effects of −0.14±0.07. Conversely, State Group 3, noted for its low inequality and moderate telehealth usage, displayed average effects of 0.06±0.04.

Figure 5 illustrates the average impact of telehealth usage across various states and all demographic categories. In the age demographic category (Fig. 5 and Fig. S2) telehealth usage demonstrated a negative impact on inequality across all state groups, with effects approximately −0.14±0.12, −0.05±0.04, and - 0.11±0.06. Regarding education and income (Figs. S3-S4). State Group 1 exhibited a negative impact of −0.06 ±0.07 and −0.16±0.14, respectively. These results validate that a decline in telehealth usage correlates with an increase in inequality, which varies by demographic categories and state.

**Figure 5.**
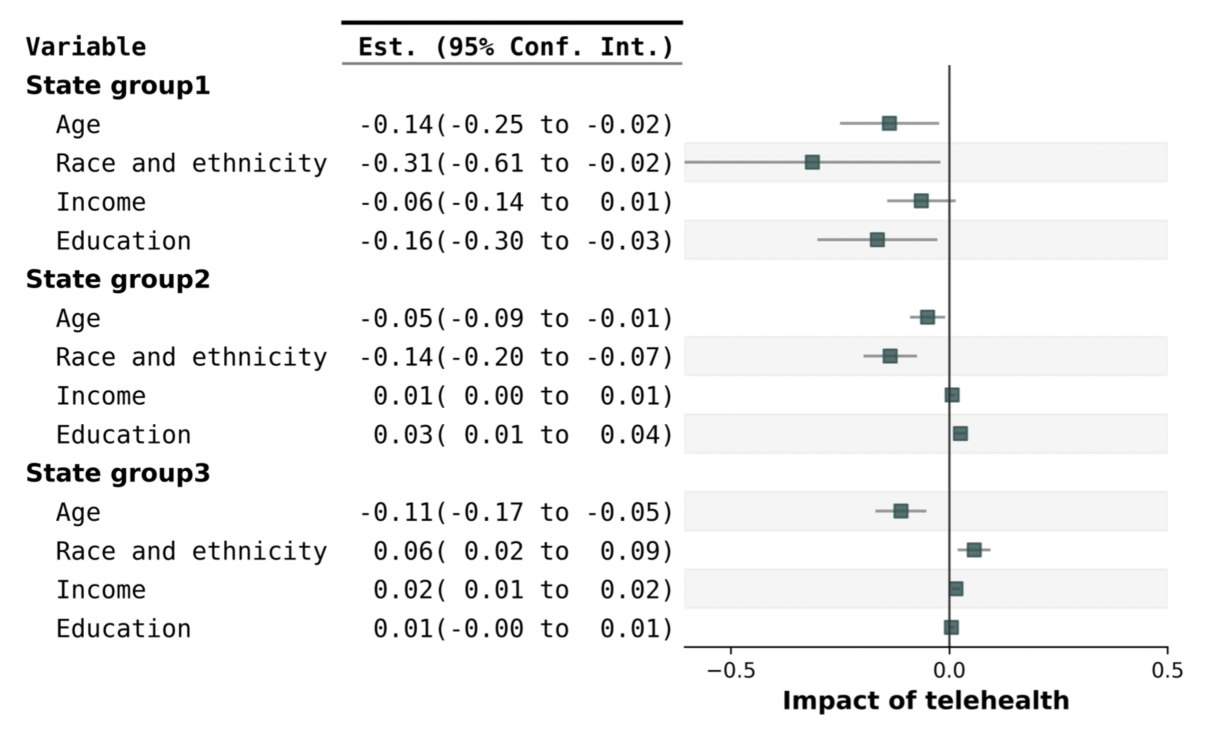
Impact of declining telehealth usage. The decline in telehealth usage has impacted healthcare inequality across different demographics. This impact is negative for all demographics for state group 1. However, in State Groups 2 and 3, the negative effects are confined to specific age groups or racial and ethnic group.

## Discussion

### Summary of findings

This study provides a comprehensive analysis of healthcare utilization during the COVID-19 pandemic across various US states. Our findings show that the pandemic considerably disrupted healthcare services across all demographics, with notable impacts in medical specialties such as emergency medicine, anesthesiology, and cardiology. By using the Gini coefficient, our analysis shows that the decline in healthcare utilization was unevenly distributed, disproportionately affecting populations with younger age, lower income, and lower education levels and the Hispanic population. Compared to the pre-pandemic period, the pandemic has markedly intensified health inequalities, showing increases of 14.5%, 2.3%, 7.0%, and 4.8% in inequalities for the demographic categories of age, race and ethnicity, income, and education, respectively. The inequality persisted throughout the pandemic, becoming more pronounced in 2022 compared to 2021, indicating the enduring impact of the long COVID-19 pandemic.

Our analysis reveals the crucial role of telehealth, the alternative to in-person healthcare visits, in mitigating disparities in healthcare access during the pandemic. Specifically, we identify that a decrease in telehealth usage is associated with heightened healthcare inequality, especially across age, race, and ethnicity demographic categories. Our state-level analysis further indicates that the distribution of healthcare utilization inequality is geographically uneven. States that exhibited pronounced pre-existing inequalities, such as New York, benefited more significantly from telehealth interventions. However, the effectiveness of telehealth in reducing these inequalities varies across different demographic groups and states.

### Limitations

While comprehensive this study is, several limitations need to be addressed in future research. First, the reliance on electronic health record datasets may introduce biases that could skew the results^39^. Future studies should consider methods to account for and mitigate these biases to improve the accuracy and reliability of the findings. Second, although the Gini coefficient is a widely accepted measure for assessing inequality, it lacks the capability to fully capture the complexities of spatial-temporal and multidimensional disparities^38^. Third, while this study focuses on telehealth as an adaptive strategy, multiple factors contribute to mitigating health inequality. More holistic approach is required to identify and evaluate the compounding efficacy of various strategies in reducing disparities^40^. In addition, the analysis does not consider the delayed or cumulative impacts of adaptive strategies, which could provide insights into long-term benefits. Lastly, the macroscopic perspective in the study may overlook critical details such as care quality, hospital bed utilization, patient-provider interactions, mental health outcomes, chronic disease management, and the implications of health policies^41–43^. Addressing these gaps in future research will enable a more detailed and nuanced understanding of health inequalities and inform more effective strategies for addressing them during pandemics.

## Conclusion

This study shows that the COVID-19 pandemic has significantly exacerbated healthcare inequalities in the United States. It highlights the critical need for targeted interventions and policy reforms to address these disparities^44,45^. This includes not only expanding telehealth but also ensuring that it is accessible and effective across all populations, coupled with broader structural changes to healthcare delivery and funding to enhance healthcare resilience against future crises.

## Acknowledgments

The data, technology, and services used in generating these research findings were generously supplied pro bono by the COVID-19 Research Database partners, who are acknowledged at https://covid19researchdatabase.org/. We acknowledge the support of Research Accelerator grants funded by the Bill & Melinda Gates Foundation.

## Data availability

The data that support the findings of this study are available from the Healthjump database and the COVID-19 Research Database consortium. However, restrictions apply to the availability of these data, which were used under license for the current study. The EMR dataset is not publicly available.

## Author Contributions

Zhong had full access to all the data in the study and take responsibility for the integrity of the data and the accuracy of the data analysis.

Concept and design: Lian, Zhong, and Gao.

Acquisition, analysis, or interpretation of data: Zhong and Lian. Drafting of the manuscript: Lian.

Critical review of the manuscript for important intellectual content: Zhong, Pei, and Gao. Administrative, technical, or material support: Zhong, Pei, and Gao.

## Supplementary Information

**Dataset.** Table S1 displays the number of patients and their visit records included in our dataset spanning from 2017 to 2022. Our analysis encompasses 17 million patients and 94 million records. We specifically focus on the three specialties with the highest patient volumes: emergency medicine, anesthesiology, and cardiology.

**Impact of telehealth.** For age, as depicted in Fig.S2, our findings indicate that 33 states experienced negative impacts due to decreased use of telehealth, with an average impact of −0.09 ±0.03. In terms of education and income, as shown in Fig. S3 and Fig. S4 respectively, 12 and 15 states exhibit negative impacts. The average impact on the education group was −0.024 ±0.034 while the effect on the impact group was notably smaller, averaging −0.004 ±0.016. This data suggests that the reduction in telehealth use has had a varied but generally negative effect on inequality across different demographic categories.

**Figure S1.**
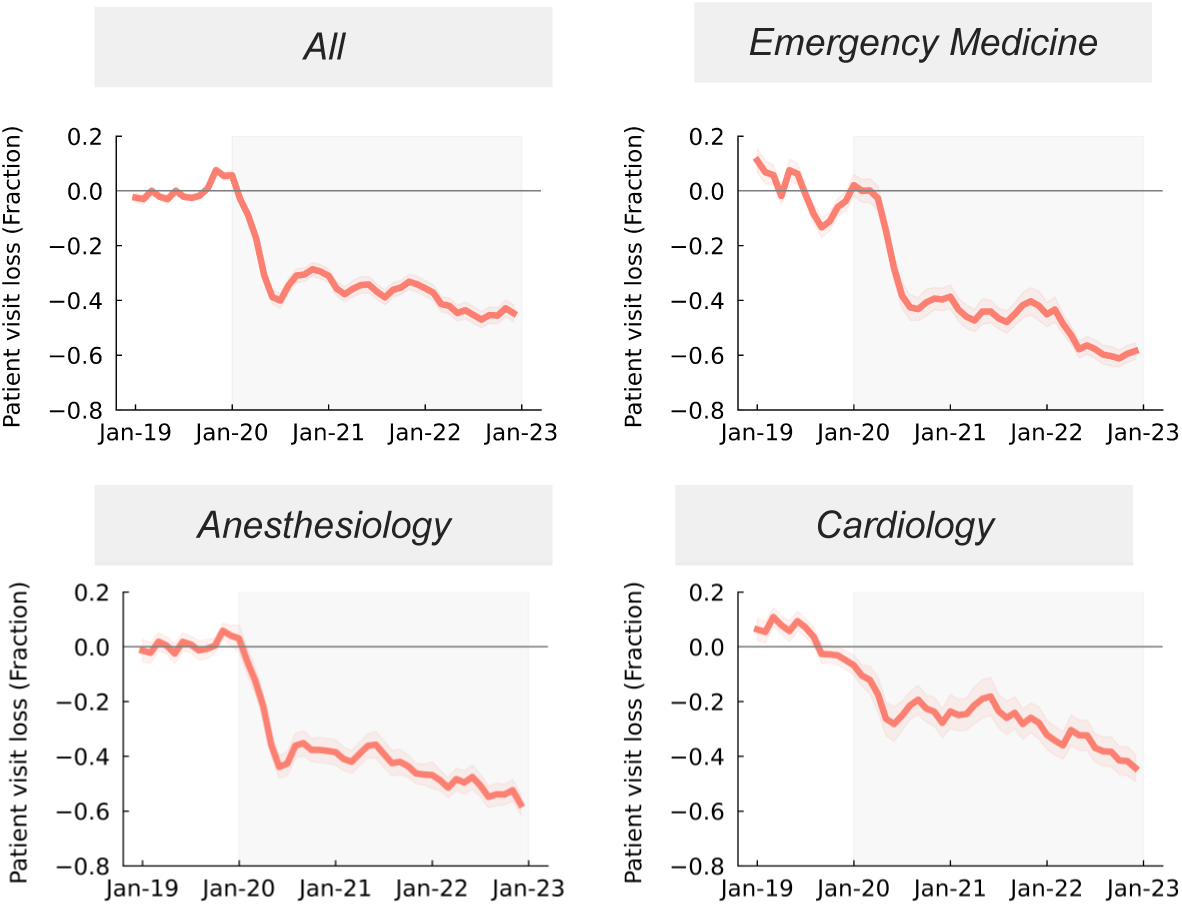
Temporal patient visit loss from 2019 to 2022.

**Figure S2.**
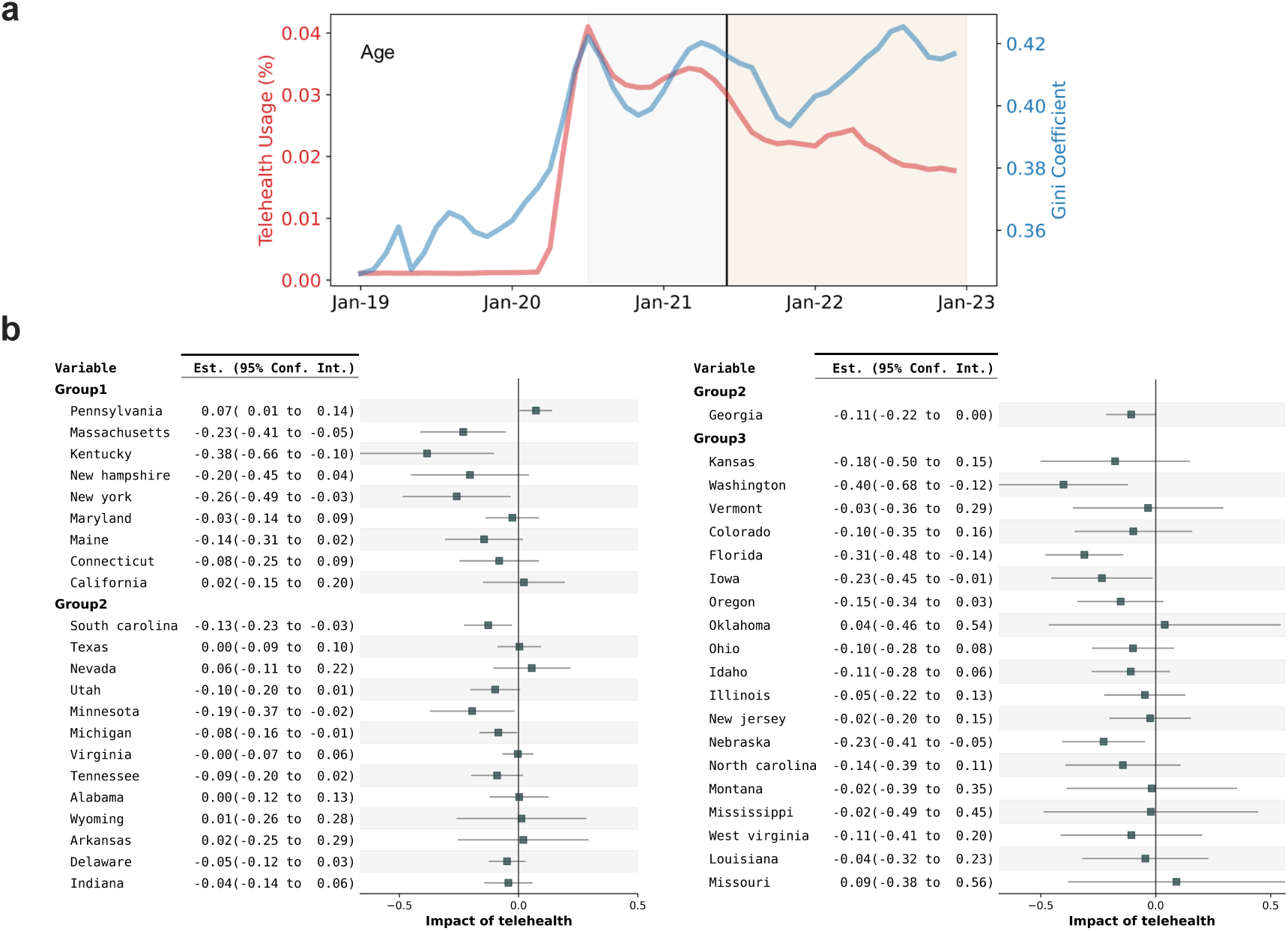
The impact of telehealth usage for age groups across states.

**Figure S3.**
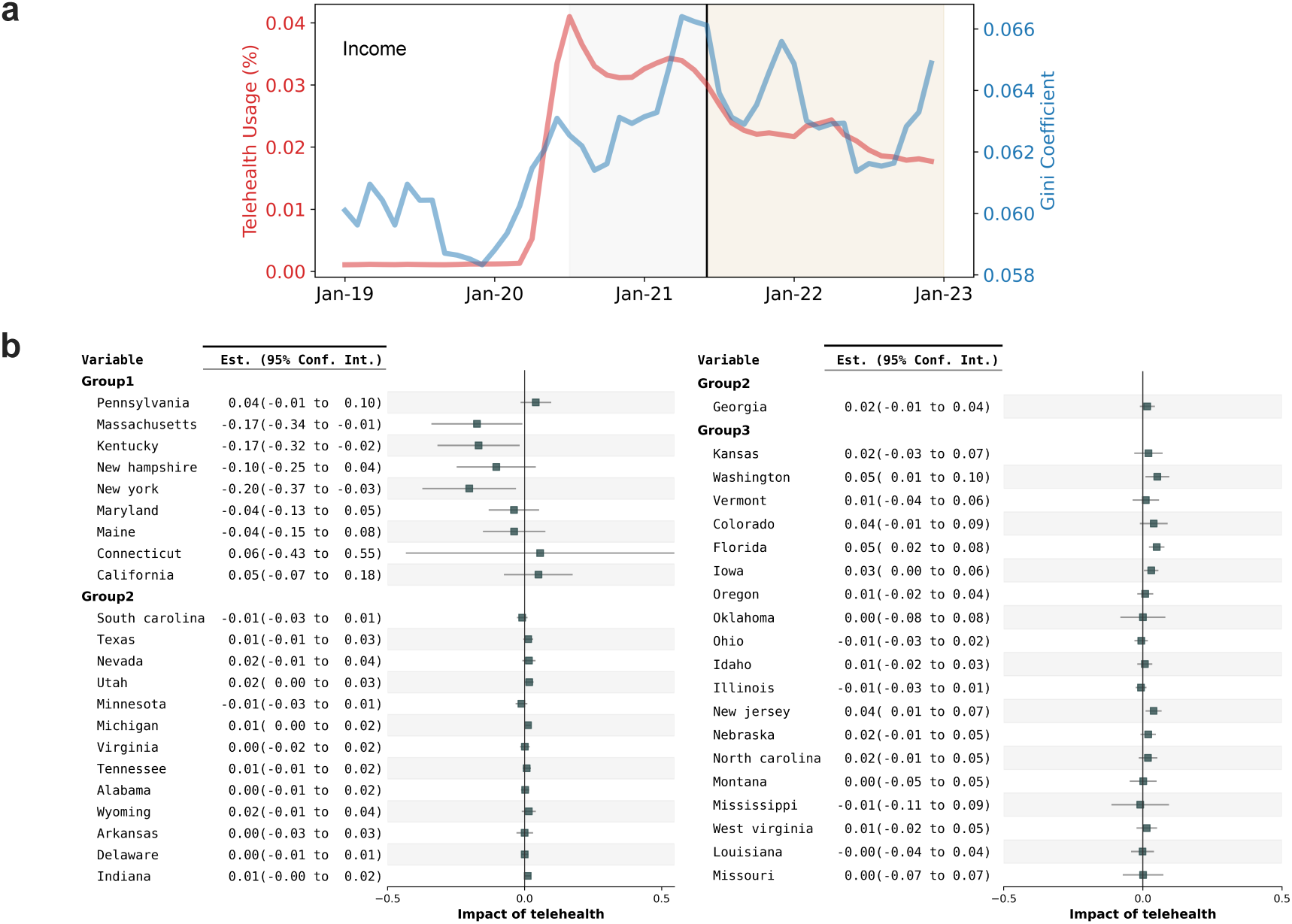
The impact of telehealth usage for income group across states.

**Figure S4.**
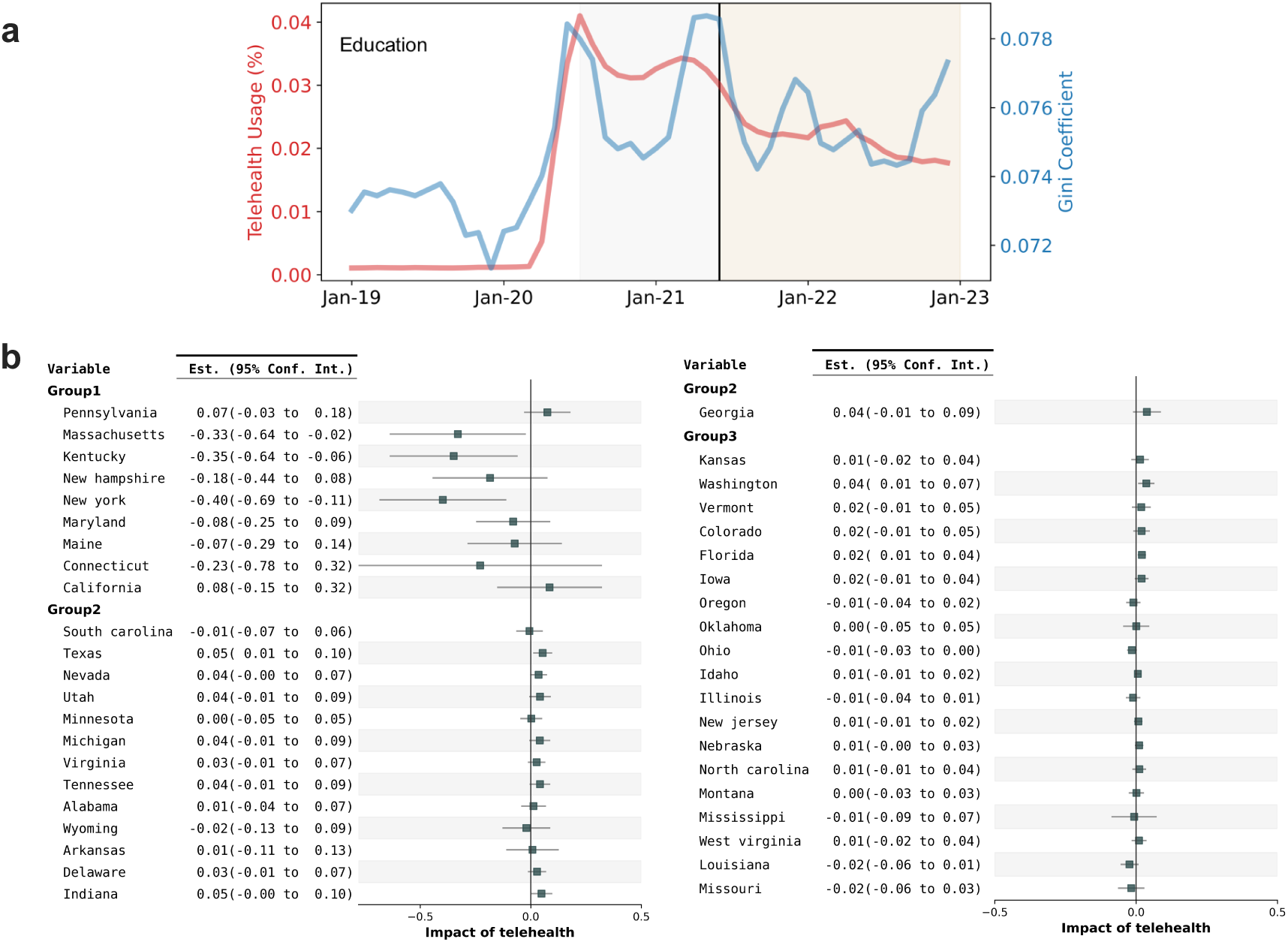
The impact of telehealth usage for education groups across states.

**Table S1.**
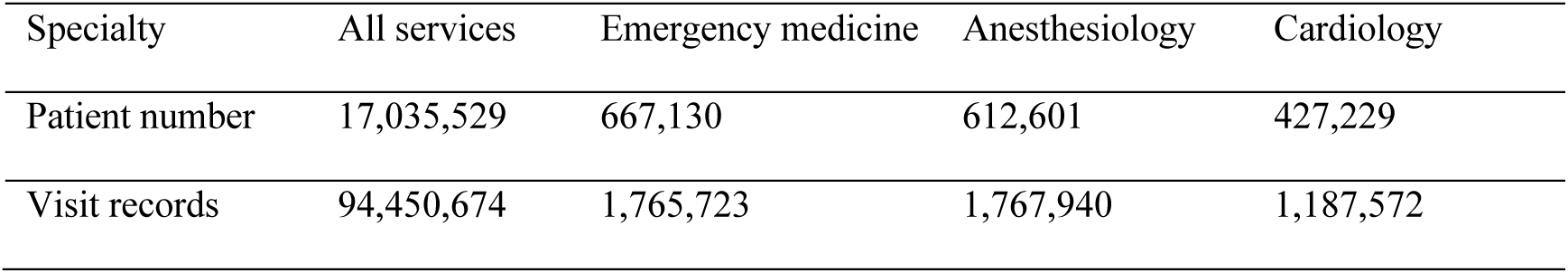
Summary of dataset.

| Specialty | All services | Emergency medicine | Anesthesiology | Cardiology |
| --- | --- | --- | --- | --- |
| Patient number | 17,035,529 | 667,130 | 612,601 | 427,229 |
| Visit records | 94,450,674 | 1,765,723 | 1,767,940 | 1,187,572 |

